# Elementary spatial structures and dispersion of COVID-19: health geography directing responses to public health emergency in São Paulo State, Brazil

**DOI:** 10.1101/2020.04.26.20080895

**Authors:** Carlos Magno Castelo Branco Fortaleza, Raul Borges Guimarães, Rafael de Castro Catão, Cláudia Pio Ferreira, Gabriel Berg de Almeida, Edmur Pugliesi

**Affiliations:** Department of Infectious Diseases. Botucatu Medical School, São Paulo State University (UNESP), Botucatu, São Paulo State, Brazil; Deparment of Geography, Faculty of Science and Technology, São Paulo State University (UNESP), Presidente Prudente, São Paulo State, Brazil; Department of Geography, Federal University of Espírito Santo, Vitória, Espírito Santo State, Brazil; Institute of Biosciences, São Paulo State University (UNESP), Botucatu, São Paulo State, Brazil

**Author notes:** These authors contributed equally to this work (Carlos Magno Castelo Branco Fortaleza, Raul Borges Guimarães). These authors also contributed equally to this work (Rafael de Castro Catão, Cláudia Pio Ferreira, Gabriel Berg de Almeida, Edmur Pugliesi).

## Abstract

Public health policies to contain the spread of COVID-19 rely mainly on non-pharmacological measures. Those measures, especially social distancing, are a challenge for developing countries, such as Brazil. In São Paulo, the most populous state in Brazil (45 million inhabitants), most COVID-19 cases up to April 18th were reported in the Capital and metropolitan area. However, the inner municipalities, where 20 million people live, are also at risk. As governmental authorities discuss the loosening of measures for restricting population mobility, it is urgent to analyze the routes of dispersion of COVID-19 in those municipalities. In this ecological study, we use geographical models of population mobility as patterns for spread of SARS-Cov-2 infection. Based on surveillance data, we identify two patterns: one by contiguous diffusion from the capital metropolitan area and other that is hierarchical, with long-distance spread through major highways to cities of regional relevance. We also modelled the impact of social distancing strategies in the most relevant cities, and estimated a beneficial effect in each and every setting studied. This acknowledgement can provide real-time responses to support public health strategies.

## Introduction

The International Health Regulations (IHR), administered by World Health Organization (WHO), was last revised in 2005, under the influence of the global response to the SARS emergency and of the risk of the H5N1 influenza pandemic [1]. Since then, it has guided coordinated international cooperation during public health emergencies such as Zika virus and Ebola epidemics [2]. However, the current COVID-19 pandemic is the greatest challenge faced by IHR thus far [3]. Although, the WHO has issued several guidelines related to the current epidemic, the level of adherence varies among nations and, inside nations, provinces and states [4].

Up to the present day, non-pharmacological interventions, like social distancing, radical lockdown and extensive testing for SARS-Cov-2 infection, have been applied by different countries, with widely varying degrees of success [5,6]. In some countries, such as Brazil, scientific research on the effectiveness of those strategies have been severely hampered by political bias, which interferes with public health decisions [7].

São Paulo, Brazil’s most populous State (45 million inhabitants), is also the most severely affected by COVID-19. The State governor has challenged Brazil’s President denialism of the pandemic, and declared closure of commerce, schools and other non-essential services. However, despite ferocious spread of the virus on the State Capital and metropolitan area, the slowly evolving of the epidemic in the inner cities of the state, where 20 million people live, has led to protests against governmental measures. In this context, there is a sense of urgency about predicting routes of spreading of the epidemic in the inner State and the risks for the population that resides there.

Here, we discussed a detailed analysis of the spatial diffusion of COVID-19 in São Paulo State, Brazil, with the objective of providing real-time responses to support public health strategies. This approach can be done in other states of Brazil as well as in other developing countries [8].

## Methods

### Geographical data modeling

Spatial analysis of surveillance data includes exploratory data analysis, spatial modelling and visualization [9]. The first one uses spatial statistical methods to measure centrality and dispersion of data sets in order to detect spatial patterns and to examine relationships between variables of the complex phenomenon under investigation. The second one examines the elementary forms of spatial organization that explains the phenomenon under study, such as railways, land cover, demographic and also social factors [10]. Lastly, visualization provides a synthesis of the previous procedures, aiming the elaboration of a thematic map that can be presented to managers for decision making in emergency situations in public health.

Focusing in the State of São Paulo, its center and periphery structure, main roads and network structure that gives population and trade mobility, the geographic spread of coronavirus was studied. For this, the date of the first confirmed case in each municipality in São Paulo State were centered at the city hall location. The six nearest neighbors (with reported cases) of each point so were used in the interpolator; and the contribution of each one was weighted by the inverse of its distance. Therefore, assuming that the measured values closest to the prediction location have more influence on the predicted value those far away, the following equation was used

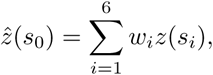

where 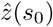, *w_i_* and *z*(*s_i_*) are the estimated value at position s_0_, the weight attributed to each pair of coordinates (1/|*s_i_* — *s*_0_|) and the number of cases observed at position *s_i_*. The gradient maps were constructed using surveillance data (number of confirmed cases of coronavirus) updated on Abril 15th, 2020.

Out of 645 municipalities in São Paulo State, 145 have confirmed cases and were used in the study. The interpolator created a surface on which the values from points (municipalities) are combined and recorded in a data matrix, simplifying information and creating regional patterns. As it has spatiotemporal data, it must be read with the darkest data in the red palette as the oldest that passes through the orange, yellow going to the blue palette, which are the municipalities that were later infected.

In the second step, data about each municipalities such as infrastructure, facilities, land use, jobs, and urban mobility were used to identify the fundamental entities of the spatial structure that triggers coronavirus dispersion in São Paulo territory [11]. In the visualization step, we attempted to produce a map that could be understandable by health authorities and community [8,9]. The proportional symbol maps scale was used to size the circles proportionally to the number of confirmed cases in each municipality. Standard deviation ellipses were drawn to show, at different times, the main direction of disease spreading. Our maps were constructed based on principles of graphic semiology, theory of colors, and visual communication [9, 12, 13].

### Epidemic modelling

A deterministic age structured model splits the human population into age groups, from 0 to 4 years, 5 years interval from 5 to 70 years, and greather that 70 years. The variables of the model are *t*, *S_i_*: = *S_i_*.(*t*), *E_i_*: = *E_i_*(*t*), *I_i_*: = *I_i_*(*t*), *R_i_*: = *R_i_* (*t*); respectively, time, susceptibles, exposed, infected, and recovered individuals. The index *i* takes into account the age class. The natural mortality rate *μ* appers in all age classes, and the parameter *α_i_* deals with transition among age classes. Individuals born susceptible, and become exposed after contacting infected individuals at rate *β*. The parameter *c_i,j_* represents the fraction of daily contacts between individuals at age group *i* and *j* [14]. The parameter ξ ∈ [0,1] takes into account the effect of social distancing. After a period of time *η*^−1^ exposed individuals becomes infectious. Additional mortality related to the disease is considered in the compartiments of infected individuals, *σ_i_*, and it is calculated through the expression

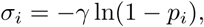

where *p_i_* is the probability that an individual at age group *i* dies during his infectious period. We have *p*_1_ = *p*_2_ = 0, *p*_3_ = … = *p*_8_ = 0.002, *p*_9_ = *p_10_* = 0.004, *p*_11_ = *p*_12_ = 0.013, *p*_13_ = *p*_14_ = 0.036, and *p*_15_ is weighted by the number of individuals in age group with more than 80 years old (*x*_1_) and in the age group between 75 to 79 years old (*x*_2_) giving

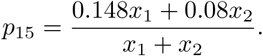

Finally, infected individuals become recovered at rate *γ*. The ordinary differential system is written in terms of population density

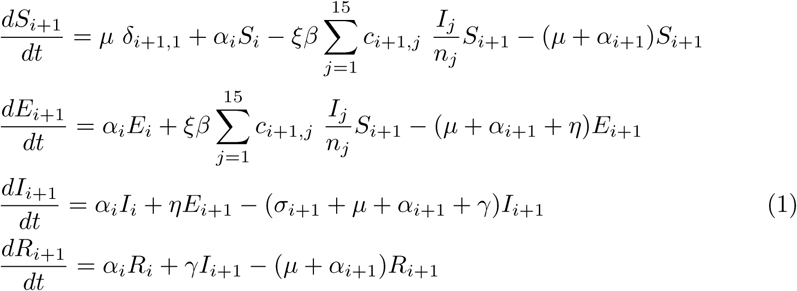

with *i* = 0,…, 14, *α*_0_ = *α*15 = 0, and *α_j_* = *α*, *j* ≠ {0,15}, *δ*1,1 = 1, and *δ_j_*_,1_ = 0, *j* ≠ 1. Besides, *n_j_* = *S_j_* + *E_j_* + *I_j_* + *R_j_*. Table 1 summarizes model parameters, their description, range of values and units.

**Table 1.** Parameters of the model, their descritption and values.

| Parameter | Description | Value |
| --- | --- | --- |
| $\mu$ | mortality rate | $1/75 \text{ years}^{-1}$ |
| $\alpha$ | transition rate among age classes | $1/5 \text{ years}^{-1}$ |
| $\sigma$ | additional mortality rate | $[0, 0.02] \text{ days}^{-1}$ |
| $\beta$ | infection rate | $0.44 \text{ days}^{-1}$ |
| $\eta^{-1}$ | latent period | 3 days |
| $\gamma^{-1}$ | infectious period | 6.4 days |
| $\xi$ | reduction on the contact rate | 0.5 |

Disease control is modelled as a reduction of 50% of the contact matrix and starts at *t* = 0 [15,16]. In all cases *R*_0_ ≈ 2.7. The simulations start with ten infected individuals (in the age class of 25 to 50 years) introduced in a whole susceptible population.

## Results and Discussion

The exploratory analysis of data on confirmed COVID-19 cases in São Paulo State generated a diffusion map in which a color spectrum indicates de areas ranging from the earlier to more recent introduction of SARS-Cov-2 (Fig 1).

**Fig 1.**
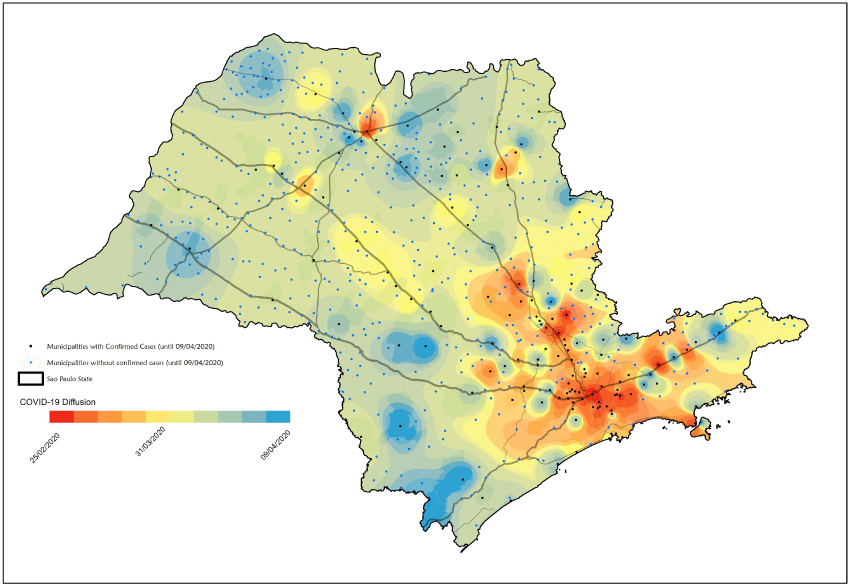
Diffusion map for COVID-19 in São Paulo State Brazil, up to April 18, 2020. The color spectrum indicates the areas of early (in red) to those of more recent COVID-19 introduction (in blue).

Based on the results of the exploratory analysis and previous population mobility studies in the State of São Paulo, two dispersion patterns were postulated. In the first one, virus dispersion occurs by contiguity, from a region of initial introduction, that is the Metropolitan Region of the Capital, the City of São Paulo (contagious diffusion) to it nearest neighborhoods. In the second one, there is a long-distance dispersion following structural axes (highways) that connect São Paulo city to pole municipalities of regional importance (hierarchical diffusion). From these, diffusion by contiguity occurs again to smaller municipalities. Fig 2 presents the diffusion axes, while Fig 3 shows the municipalities of regional relevance for the disease spread.

**Fig 2.**
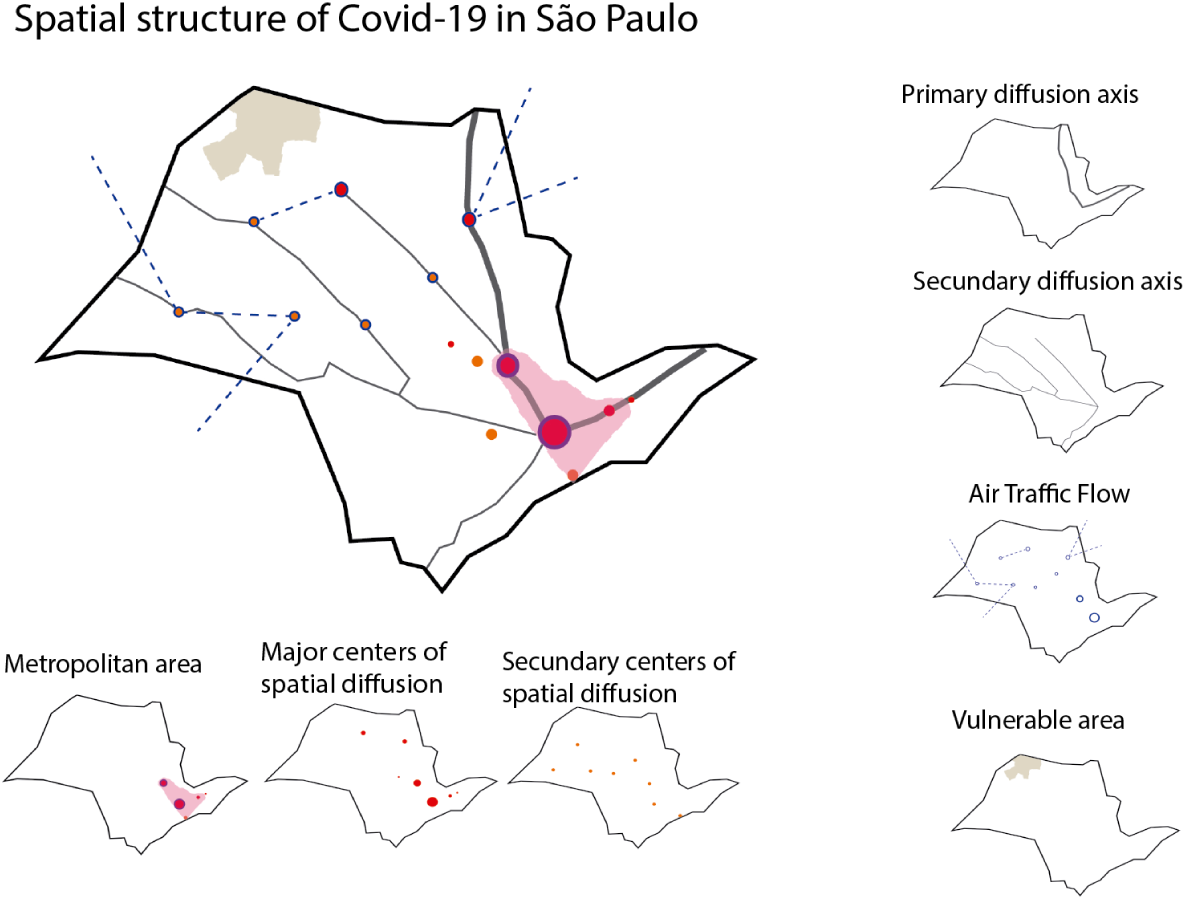
Elementary spatial structures associated to COVID-19 spread in Sâo Paulo State, Brazil.

**Fig 3.**
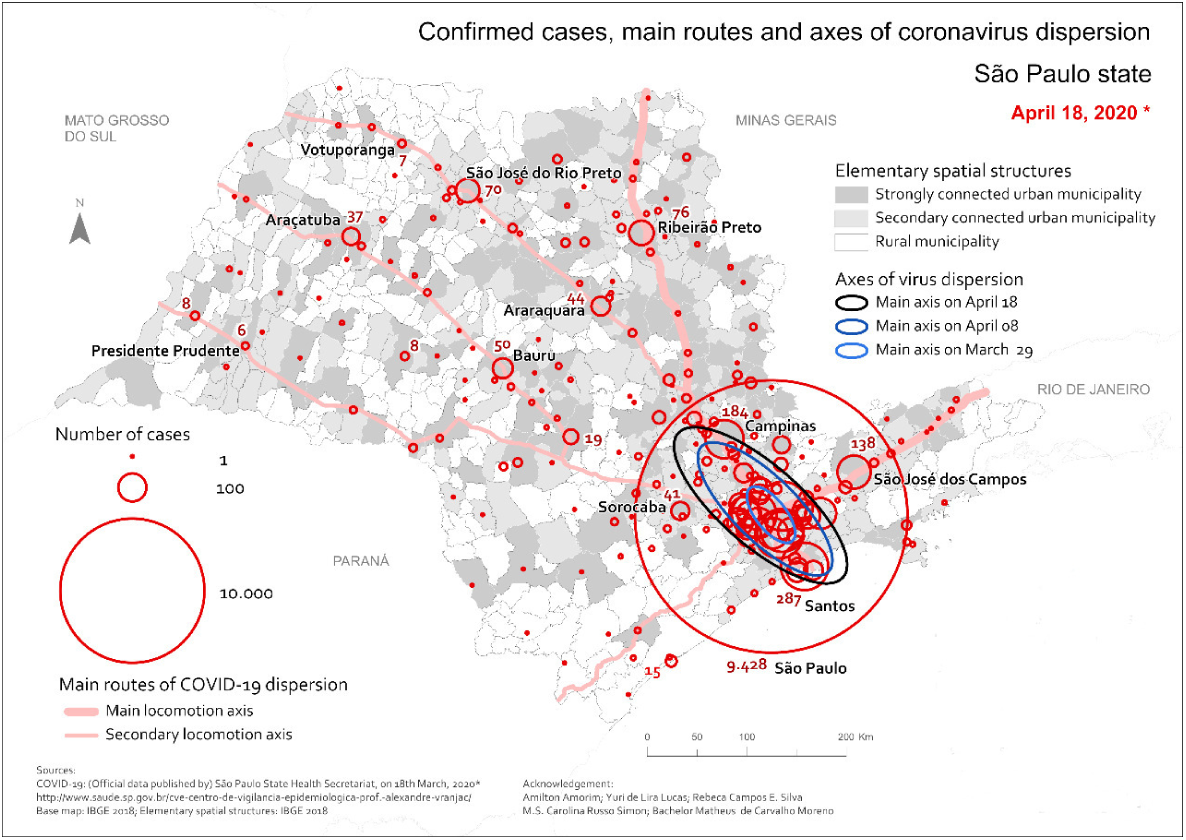
Distribution of confirmed COVID-19 cases in São Paulo State as of April 18th 2020, Brazil, with identification of municipalities that influence regional spread.

In the next step, we simulated a susceptible-exposed-infected-recover (SEIR) model for each city of regional influence, with or without social distancing measures [16, 17]. The results are shown in Figs 4 and 5. Demographic characteristics and number of confirmed COVID-19 cases (up to April 18, 2020) are presented in Table 2.

**Fig 4.**
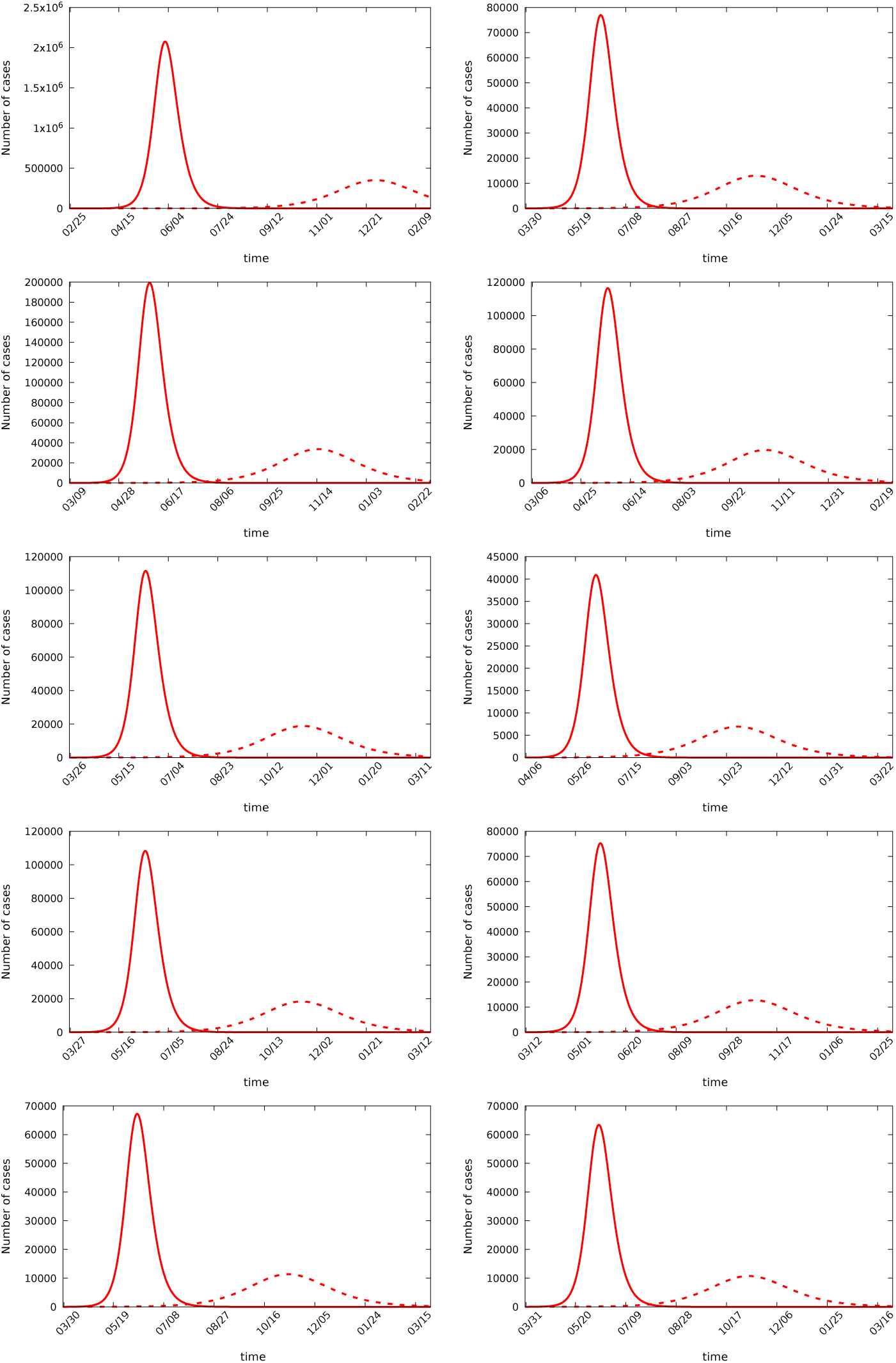
Temporal evolution of the epidemics. From top to bottom and left to right we have São Paulo, Santos, Campinas, São José dos Campos, Ribeirão Preto, São Carlos, Sorocaba, São José do Rio Preto, Piraciaba and Bauru. The continuous and dashed lines correspond to the case without control and with control.

**Fig 5.**
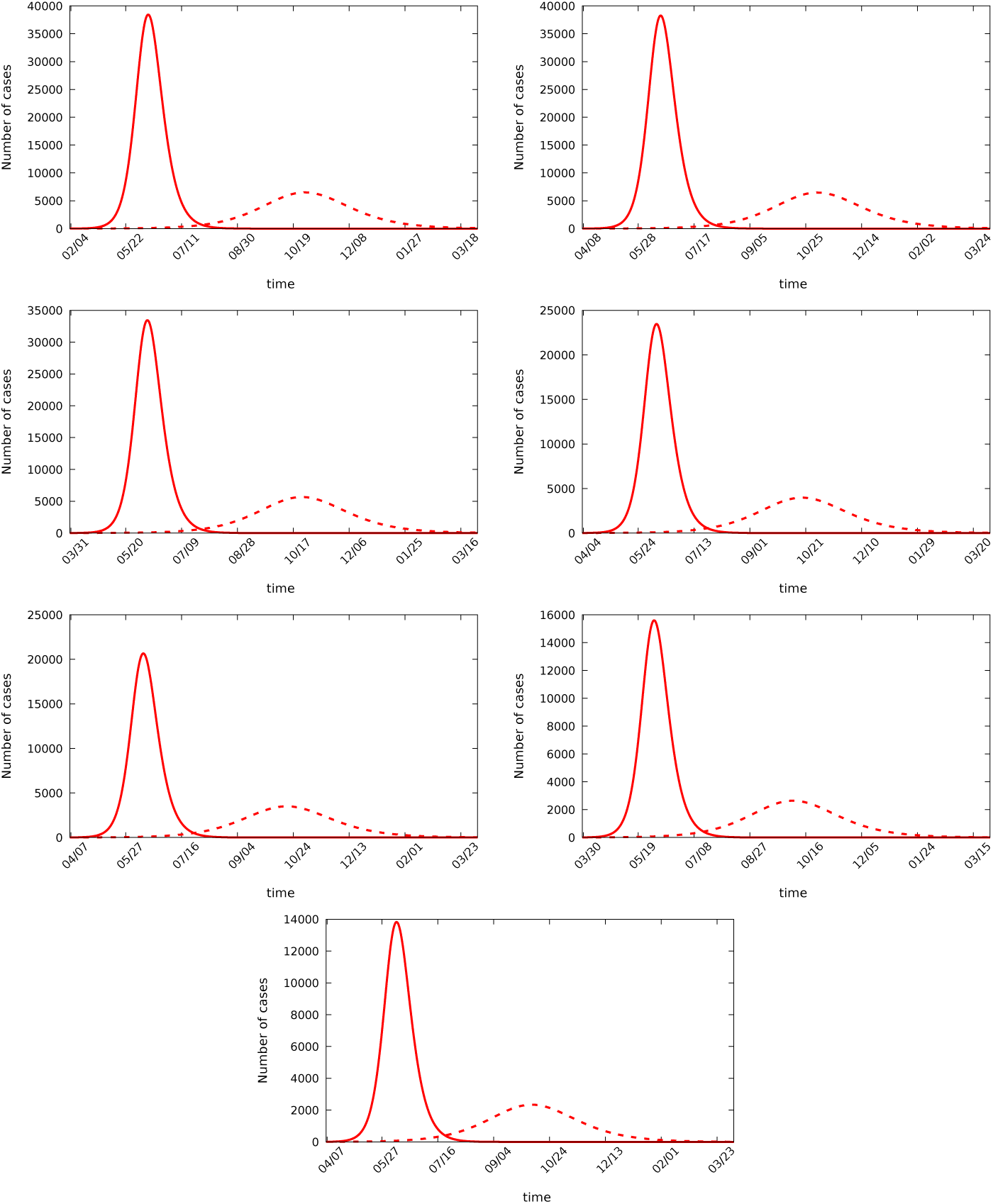
Temporal evolution of the epidemics. From top to bottom and left to right we have Araraquara, Presidente Prudente, Araçatuba, Botucatu, Barretos, Votuporanga and Bebedouro. The continuous and dashed lines correspond to the case without control and with control.

**Table 2.** Epidemiologic COVID-19 data for São Paulo State capital and cities of regional importance.

| **Municipality** | **Population**<br>**(inhabitant)** | **Dist.<sup>1</sup>**<br>**(Km)** | **Connection with**<br>**the Capital<sup>2</sup>** | **Cumul.**<br>**cases** | **Incid.<sup>3</sup>** | **Cumul.**<br>**deaths** | **Mort.<sup>3</sup>** | **SRD** | **SRD<sup>3</sup>**<br>**incid.** |
| --- | --- | --- | --- | --- | --- | --- | --- | --- | --- |
| São Paulo (capital) | 12252023 | - | - | 9815 | 80.11 | 715 | 5.84 | 14078 | 114.90 |
| Santos | 433311 | 55 | Contiguity | 310 | 71.54 | 19 | 4.38 | 209 | 48.23 |
| Campinas | 1204073 | 95 | Contiguity | 192 | 15.95 | 7 | 0.58 | 652 | 54.15 |
| São J. dos Campos | 721944 | 91 | Contiguity | 136 | 18.84 | 3 | 0.42 | 301 | 41.69 |
| Ribeirão Preto | 703293 | 314 | Primary axis | 81 | 11.52 | 5 | 0.71 | 323 | 45.93 |
| São Carlos | 251983 | 231 | Primary axis | 9 | 3.57 | 2 | 0.79 | 138 | 54.77 |
| Sorocaba | 679378 | 100 | Secondary axis | 50 | 7.36 | 9 | 1.32 | 216 | 31.79 |
| São J. do Rio Preto | 460671 | 440 | Secondary axis | 71 | 15.41 | 7 | 1.52 | 268 | 58.18 |
| Piracicaba | 404142 | 162 | Secondary axis | 21 | 5.20 | 2 | 0.49 | 124 | 30.68 |
| Bauru | 376818 | 343 | Secondary axis | 58 | 15.39 | 3 | 0.80 | 127 | 33.70 |
| Araraquara | 236072 | 273 | Secondary axis | 49 | 20.76 | 2 | 0.85 | 74 | 31.35 |
| Presidente Prudente | 228743 | 550 | Secondary axis | 6 | 2.62 | 2 | 0.87 | 47 | 20.55 |
| Araçatuba | 197016 | 530 | Secondary axis | 39 | 19.80 | 0 | 0.00 | 45 | 22.84 |
| Botucatu | 146497 | 230 | Secondary axis | 27 | 18.43 | 2 | 1.37 | 70 | 47.78 |
| Barretos | 122098 | 424 | Secondary axis | 9 | 7.37 | 0 | 0.00 | 50 | 40.95 |
| Votuporanga | 94547 | 519 | Secondary axis | 9 | 9.52 | 0 | 0.00 | 42 | 44.42 |
| Bebedouro | 77496 | 384 | Secondary axis | 2 | 2.58 | 0 | 0.00 | 45 | 58.07 |
| Registro | 56322 | 175 | Secondary axis | 4 | 7.10 | 1 | 1.78 | 25 | 44.39 |

1. distance from the capital; 2. classification according to Fig 3; 3. incidence or mortality per 100,000 inhabitants; 3. number of hospital admissions for severe respiratory disease (SRD).

As one can infer from Fig 3 and Table 2, there is great heterogeneity in inner São Paulo State, and some cities have special relevance due to their geographic localization, either proximity to São Paulo City or laying alongside high mobility highways. Simulations (Figs 4 and 5) show that, in those municipalities, social distancing measures can mitigate the impact of pandemics in a way that is much similar to what has been demonstrated for São Paulo City metropolitan area [18, 19]. We can also note that Campinas, São José dos Campos and Santos, all contiguity to São Paulo city, are strongly affected by the spatio-temporal evolution of the disease at the metropolitan area of São Paulo, while rural municipalities are less affected. Santos, that has a huge mortality per 100,000 inhabitants, is the one in the list which has the biggest number of older population (≥ 50 years).

Though our research has the limitations inherent to the ecological study design [11], our predictions of routes and risks of COVID-19 in inner São Paulo State (Fig 2) have been thus far validated by surveillance data (Fig 3). Given the extensive mobility between smaller municipalities and those cities with regional economic relevance [11], it is reasonable to infer that the regional spread of SARS-Cov-2 infections depends on the success of non-pharmacological strategies applied in the latter.

São Paulo State distancing measures started on March 22nd and is presently under heavy pressure from several sectors of industry and trading companies. As the local government hints at the possibility of loosening restrictive measures, it is urgent to provide a way of protecting the population health. We also state that similar methodological approaches can direct public health strategies in other developing countries, especially those that either have great territorial extension and/or have diverse patterns of urbanization and mobility.

## Conclusion

Spatial analysis of coronavirus spread can provide are an important tool for public health management, highlighting the routes of disease dispersion and the fragility of municipalities related to its socio-demographic characteristics. The main routes of dispersion from Capital to inner State are the hallways that give mobility to people and merchandise. Currently, non-pharmacological controls are the only tools to control disease spreading among individuals and municipalities. The existence of two different ways of disease dispersal, by standard diffusion and hierarchical one can provide alternative strategies to control disease spread.

## Data Availability

Data will be available upon contact to the corresponding author

## Acknowledgments

We thank to Centro de Vigilância Epidemiológica of São Paulo State (CVE), Health Department, to provide the data. CPF also thanks support f rom São Paulo Research Foundation (FAPESP) grant 18/24058–1.

